# Clinical manifestations of patients with Coronavirus Disease 2019 (COVID-19) attending at hospitals in Bangladesh

**DOI:** 10.1101/2020.07.30.20165100

**Authors:** Md. Shahed Morshed, Abdullah Al Mosabbir, Prodipta Chowdhury, Sheikh Mohammad Ashadullah, Mohammad Sorowar Hossain

**Author notes:** Corresponding author: Mohammad Sorowar Hossain, Biomedical Research Foundation, Dhaka, Bangladesh.

## Abstract

Bangladesh is in the rising phase of the ongoing pandemic of the coronavirus disease 2019 (COVID-19), caused by severe acute respiratory syndrome coronavirus-2 (SARS-CoV-2). The scientific literature on clinical manifestations of COVID-19 patients from Bangladesh is scarce. This study aimed to report the sociodemographic and clinical characteristics of patients with COVID-19 in Bangladesh. We conducted a cross-sectional study at three dedicated COVID-19 hospitals. The severity of the COVID-19 cases was assessed based on the WHO interim guidance. Data were collected only from non-critical COVID-19 patients as critical patients required immediate intensive care admission making them unable to respond to the questions. A total of 103 RT-PCR confirmed non-critical COVID-19 patients were enrolled. Most of the patients (71.8%) were male. Mild, moderate and severe illness were assessed in 74.76%, 9.71% and 15.53% of patients respectively. Nearly 52.4% of patients had a co-morbidity, with hypertension being the most common (34%), followed by diabetes mellitus (21.4%) and ischemic heart disease (9.7%). Fever (78.6%), weakness (68%) and cough (44.7%) were the most common clinical manifestations. Other common symptoms included loss of appetite (37.9%), difficulty in breathing (37.9%), altered sensation of taste or smell (35.0%), headache (32%) and body ache (32%). The median time from onset of symptom to attending hospitals was 7 days (IQR 4-10). This study will help both the clinicians and epidemiologists to understand the magnitude and clinical spectrum of COVID-19 patients in Bangladesh.

## Introduction

The world has been experiencing one of the most serious public health crises in the history of humankind. The ongoing pandemic of coronavirus disease 2019 (COVID-19) is caused by severe acute respiratory syndrome coronavirus-2 (SARS-CoV-2). As of 28 July 2020, over 16 million individuals have been infected with over 650,000 deaths worldwide^1^. SARS-COV-2 infection predominantly results in an acute respiratory illness. In addition, it can cause a myriad of extrapulmonary symptoms. The clinical spectrum ranges from asymptomatic or mildly symptomatic flu-like illness to potentially life-threatening critical conditions^2^. Recent studies suggest that the clinical spectrum of COVID-19 can vary among different ethnicities and geographical locations across the world^3^.

Owing to a population of over 160 million, inadequate healthcare system, and poor personal hygiene among the general population, Bangladesh is considered one of the high-risk countries for coronavirus spread. The first official case of COVID-19 was reported on 8 March 2020, and the epidemic still appears to be in a growing phase. As of 30 July 2020, a total of 234,889 cases and 3083 deaths have been reported in Bangladesh^1^. However, the information on clinical manifestations from Bangladesh is scarce in the literature. Therefore, this study aimed to document the clinical spectrum of COVID-19 patients attending fever clinics in Bangladesh.

## Methods

This was a cross-sectional study conducted among RT-PCR confirmed COVID-19 patients attending the fever clinic of a dedicated COVID-19 Hospital (Kurmitola general hospital) in Dhaka city of Bangladesh and two Upazila health complexes from different districts (Jessore and Jhenaidah) from 5 July to 18 July 2020. Diagnosis of SARS-COV-2 infection and assessment of severity were done based on the WHO interim guidance^4^. Data were collected only from non-critical COVID-19 patients as critical patients required immediate intensive care admission making them unable to respond to the questions. Socio-demographic and clinical data were evaluated and collected by experienced clinicians using a pretested case record form. Verbal consent was taken from all participants. The institutional review board of Biomedical Research Foundation, Bangladesh approved the study protocol (Ref. no: BRF/ERB/2020/003).

Data were analysed by SPSS Statistics software 22.0 (Armonk, NY: IBM Corp). Data were expressed in number (percentages, %) and median (interquartile range, IQR) as appropriate. We used Pearson’s chi square test, Fisher’s exact test and Kruskal Wallis test to compare differences between mild, moderate and severe patients where appropriate. Logistic regression was used to study associations. P <0.5 was set as statistically significant.

## Results

A total of 103 laboratory-confirmed COVID-19 patients were enrolled. About 75% (77) of cases presented with mild symptoms, followed by nearly 15% severe and 10% moderate cases. Overall, the median age of the participants was 37 years (IQR: 31-53); more than 80% of these patients were under 60 years (Table 1). Most of the patients were male (71.8%). More than half of the patients (52.4%) had at least one co-morbidity, including hypertension in 35 (34%), diabetes mellitus in 22 (21.4%) and ischaemic heart disease in 10 (9.7%) patients. Notably, around 80% of moderate and severe cases had comorbidity. The median time from onset of symptom to attending fever clinic was 7 days (IQR 4-10).

**Table 1.** Characteristics of patients with COVID-19 (N= 103)

| Variables | Severity of patients with COVID-19 |  |  |  | p |
| --- | --- | --- | --- | --- | --- |
|  | Total<br>n (%) | Mild | Moderate | Severe |  |
| Patients' number | 103 (100) | 77 (74.76) | 10 (9.71) | 16 (15.53) |  |
| Age (years), median (IQR) | 37.0<br>(31.0, 53.0) | 35.0<br>(29.0, 45.0) | 52.5 (38.0,<br>63.25) | 62.5 (42.5,<br>65.0) | **<0.001\*** |
| Age group |  |  |  |  |  |
| ≤60 years | 84 (81.6) | 70 (90.9) | 7 (70.0) | 7 (43.8) | **<0.001** |
| >60 years | 19 (18.4) | 7 (9.1) | 3 (30.0) | 9 (56.3) |  |
| Gender |  |  |  |  |  |
| Male | 74 (71.8) | 55 (71.4) | 9 (90.0) | 10 (62.5) | 0.38 |
| Female | 29 (28.2) | 22 (28.6) | 1 (10.0) | 6 (37.5) |  |
| Area of residence |  |  |  |  |  |
| Urban | 69 (67) | 51 (66.2) | 6 (60.0) | 12 (75.0) | 0.73 |
| Rural | 34 (33.0) | 26 (33.8) | 4 (40.0) | 4 (25.0) |  |
| Smoking habit |  |  |  |  |  |
| Smokers | 32 (31.1) | 25 (32.5) | 3 (30.0) | 4 (25.0) | 0.94 |
| Nonsmokers | 71 (68.9) | 52 (67.5) | 7 (70.0) | 12 (75.0) |  |
| Comorbidities |  |  |  |  |  |
| At least one comorbidity | 54 (52.4) | 33 (42.9) | 8 (80.0) | 13 (81.3) | **0.003** |
| Hypertension | 35 (34.0) | 22 (28.6) | 4 (40.0) | 9 (56.3) | 0.09 |
| Diabetes mellitus | 22 (21.4) | 10 (13.0) | 4 (40.0) | 8 (50.0) | **0.002** |
| Ischemic heart disease | 10 (9.7) | 4 (5.2) | 2 (20.0) | 4 (25.0) | **0.02** |
| Chronic kidney disease | 8 (7.8) | 3 (3.9) | 1 (10.0) | 4 (25.0) | **0.02** |
| Bronchial asthma | 6 (5.8) | 5 (6.5) | 0 (0.0) | 1 (6.3) | 1.00 |
| Chronic obstructive<br>pulmonary disease | 3 (2.9) | 1 (1.3) | 1 (10.0) | 1 (6.3) | 0.16 |
| **Others | 16 (15.5) | 11 (14.3) | 1 (10.0) | 4 (25.0) | 0.53 |

Within parentheses are percentages over column total of respective variable
Pearson's chi square test/ Fisher's exact test was done as appropriate \*Kruskal Wallis test was done
\*\* Other comorbidities: Cerebrovascular disease 3 (2.9%), dyslipidemia 3 (2.9%), acute kidney injury 2 (1.9%), chronic liver disease 2 (1.9%), malignancy 1 (1.0%), Graves' thyrotoxicosis 1 (1.0%), nonalcoholic fatty liver disease 1 (1.0%), benign enlargement of prostate 1 (1.0%), allergic rhinitis 1 (1.0%), schizophrenia 1 (1.0%)

Overall, the most common symptoms reported were fever (78.6%), weakness (68%) and cough (44.7%) followed by loss of appetite (37.9%), difficulty in breathing (37.9%), altered sensation of taste or smell (35.0%), headache (32%) and body ache (32%). Less common symptoms included sore throat (28.2%), diarrhoea (22.3%) and chest pain (14.6%). Fever was the most prevalent symptom in all groups of patients. Interestingly, 80% of moderate patients experienced difficulty in breathing compared to 62.5% severe patients (Table 2). More than half of the severe cases had tachycardia (56.3%) and tachypnoea (56.3%) at presentation; their median oxygen saturation was 87.5% (IQR 77.25-89.0).

**Table 2.** Clinical features of patients with COVID-19 at presentations (N= 103)

| Variables | Total<br>n (%) | Severity of COVID-19 |  |  | p |
| --- | --- | --- | --- | --- | --- |
|  |  | Mild | Moderate | Severe |  |
| Onset of symptom to consultation (days), median (IQR) | 7.0 (4.0, 10.0) | 7.0 (3.0, 10.0) | 10.0 (6.75, 12.0) | 9.5 (6.25, 10.0) | 0.09* |
| Symptoms |  |  |  |  |  |
| Fever | 81 (78.6) | 57 (74.0) | 10 (100.0) | 14 (87.5) | 0.13 |
| Weakness/ fatigue | 70 (68.0) | 52 (67.5) | 6 (60.0) | 12 (75.0) | 0.73 |
| Cough | 46 (44.7) | 34 (44.2) | 5 (50.0) | 7 (43.8) | 0.95 |
| Loss of appetite | 39 (37.9) | 26 (33.8) | 7 (70.0) | 6 (37.5) | 0.10 |
| Difficult breathing | 39 (37.9) | 21 (27.3) | 8 (80.0) | 10 (62.5) | <0.001 |
| Altered smell/ taste | 36 (35.0) | 30 (39.0) | 4 (40.0) | 2 (12.5) | 0.10 |
| Headache | 33 (32.0) | 25 (32.5) | 3 (30.0) | 5 (31.3) | 1.00 |
| Bodyache | 33 (32.0) | 23 (29.9) | 5 (50.0) | 5 (31.3) | 0.43 |
| Sore throat | 29 (28.2) | 27 (35.1) | 0 (0.0) | 2 (12.5) | 0.02 |
| Diarrhea | 23 (22.3) | 18 (23.4) | 1 (10.0) | 4 (25.0) | 0.73 |
| Chest pain | 15 (14.6) | 13 (16.9) | 2 (20.0) | 0 (0.0) | 0.14 |
| Runny nose | 12 (11.7) | 9 (11.7) | 1 (10.0) | 2 (12.5) | 1.00 |
| Chill | 11 (10.7) | 8 (10.4) | 0 (0.0) | 3 (18.8) | 0.35 |
| Others | 23 (22.3) | 17 (22.1) | 1 (10.0) | 5 (31.3) | 0.43 |
| Signs at presentation |  |  |  |  |  |
| Tachycardia (pulse>100 beats/ minute) | 37 (35.9) | 26 (33.8) | 2 (20.0) | 9 (56.3) | 0.13 |
| Fever (T >38°C) | 16 (15.5) | 12 (15.6) | 1 (10.0) | 3 (18.8) | 0.90 |
| Tachypnea (respiratory rate >24 breaths/ minute) | 11 (10.7) | 0 (0.0) | 2 (20.0) | 9 (56.3) | <0.001 |
| Systolic blood pressure (mm-Hg), median (IQR) | 120 (110, 136) | 120 (110, 130) | 125 (113.5, 140) | 138 (120, 148.8) | 0.02* |
| Oxygen saturation (%), median (IQR) | 96.0 (93.0, 98.0) | 97.0 (95.0, 98.0) | 93.0 (91.75, 98.0) | 87.5 (77.25, 89.0) | <0.001* |

Within parentheses are percentages over column total of respective variable, Pearson's chi square test/ Fisher's exact test was done as appropriate \*Kruskal Wallis test was done
**Other symptoms:** Vomiting 8 (7.8%), abdominal pain 6 (5.8%), dizziness 5 (4.9%), red eye 4 (3.9%)

## Discussion

This study aimed to determine the clinical characteristics of RT-PCR confirmed noncritical patients with COVID-19 attending fever clinics of government hospitals in Bangladesh.

The most prevalent symptoms of non-critical COVID-19 patients in Bangladesh consist of fever (78.6%), fatigue (68%) and cough (44.7%). Similarly, in a meta-analysis from China, most prevalent symptoms were fever (80.4%), cough (63.1%) and fatigue (46%)^5^. However, studies from China included both critical and non-critical patients. In contrast, one study from Europe on mild to moderate patients reported that headache (70.3%), loss of smell (70.2%), nasal obstruction (67.8%) were the most common symptoms; fever was reported by only 45.4% of patients. Interestingly, 39% of mild cases, 40% of moderate cases and 12.5% of severe cases reported the altered sensation of taste or smell in this study. While olfactory and gustatory dysfunctions were prevalent symptoms in European patients, they were only rarely reported in Chinese patients^6,7^.

In this study, about 15% of cases were presented with severe symptoms. This is consistent with a summary report of 72,314 cases from China^8^. As expected, most of the severe patients (81.3%) had co-morbidity. Age >60 years, patients with diabetes mellitus, ischemic heart disease and chronic kidney disease had significantly higher odds of developing severe disease at presentation (Suppl. Table 1).

Our study has some limitations. First, the sample size of this study was small. Second, we could not include critical patients due to the requirement of emergency management. Therefore, our findings could not be generalized in the context of Bangladesh.

Our study reports the presenting symptoms of SARS-COV-2 infections among the Bangladeshi population. Although there are certain similarities in the range of symptoms with the Chinese population, where the pandemic originated, there are some unique findings like the high prevalence of olfactory and gustatory dysfunctions. This study will help both the clinicians and epidemiologists to understand the magnitude and clinical spectrum of COVID-19 patients in Bangladesh.

## Data Availability

Not applicable

## Author contribution

MSM. MSH and AAM designed the study, gave professional guidance and wrote the manuscript. MSM, PC and SMA collected and analysed data.

## Acknowledgement

We are grateful to Brigadier General Jamil Ahmad, director, Kurmitola General Hospital (KGH); Dr. Tania Easmin, medical officer, KGH; Dr. Md. Rashed Al Mamun, upazilla health and family planning officer (UHFPO), Shailkupa, Jhenaidah and Dr. Md. Alamgir, UHFPO, Keshabpur, Jessore for their generous support in data collection.

## Supplementary materials

**Suppl. Table 1.** Binary logistic regression analysis to predict the severe disease at presentation

| Covariates | OR (95% CI) | p |
| --- | --- | --- |
| Age >60 years | 9.90 (3.02, 32.45) | <0.001 |
| Diabetes mellitus | 5.21 (1.68, 16.22) | 0.004 |
| Ischemic heart disease | 4.50 (1.11, 18.30) | 0.04 |
| Chronic kidney disease | 6.92 (1.52, 31.38) | 0.01 |

CI, confidence interval; OR, odds ratio
Reference category is nonsevere illness (mild and moderate disease)

